# Estimating incidence of infection from diverse data sources: Zika virus in Puerto Rico, 2016

**DOI:** 10.1101/2020.10.14.20212134

**Authors:** Talia M. Quandelacy, Jessica M. Healy, Bradford Greening, Dania M. Rodriguez, Koo-Whang Chung, Matthew J. Kuehnert, Brad J. Biggerstaff, Emilio Dirlikov, Luis Mier-y-Teran-Romero, Tyler M. Sharp, Stephen Waterman, Michael A. Johansson

## Abstract

Emerging epidemics are challenging to track. Only a subset of cases is recognized and reported, as seen with the Zika virus (ZIKV) epidemic where large proportions of infection were asymptomatic. However, multiple imperfect indicators of infection provide an opportunity to estimate the underlying incidence of infection. We developed a modeling approach that integrates a generic Time-series Susceptible-Infected-Recovered epidemic model with assumptions about reporting biases in a Bayesian framework and applied it to the 2016 Zika epidemic in Puerto Rico using three indicators: suspected arboviral cases, suspected Zika-associated Guillain-Barré Syndrome cases, and blood bank data. Using this combination of surveillance data, we estimated the peak of the epidemic occurred during the week of August 15, 2016 (the 33^rd^ week of year), and 120 to 140 (50% credible interval [CrI], 95% CrI: 97 to 170) weekly infections per 10,000 population occurred at the peak. By the end of 2016, we estimated that approximately 890,000 (95% CrI: 660,000 to 1,100,000) individuals were infected in 2016 (26%, 95% CrI: 19% to 33%, of the population infected). Utilizing multiple indicators offers the opportunity for real-time and retrospective situational awareness to support epidemic preparedness and response.

## Introduction

The emergence and rapid spread of Zika virus (ZIKV), an arbovirus transmitted by *Aedes* species mosquitoes, in the Americas (1) resulted in large-scale epidemics throughout the tropical areas of the region. The first confirmed locally acquired ZIKV casein Puerto Rico was reported on December 31, 2015 (2), followed by more than 36,000 confirmed cases in 2016 (3). While confirmed cases provided an indicator of transmission intensity, reported cases represented a small proportion of actual infections (4) in part because many ZIKV infections are asymptomatic or mild, and are not captured by surveillance systems (5–7). Furthermore, distinguishing symptomatic (i.e. disease) cases of ZIKV infections from other arboviral infections (e.g., dengue, chikungunya) was difficult due to their similar symptoms (e.g. fever, rash), and serological cross-reactivity with dengue viruses (DENV). Despite these challenges, estimating the underlying ZIKV infection incidence was critical to assess useful metrics (e.g., transmission intensity, the number of people previously infected, and the number still at risk) that informed prevention and response measures, and preparation for severe outcomes like Guillain-Barré Syndrome (GBS) (8) and congenital Zika syndrome (9).

ZIKV serosurveys, like those conducted in Yap and French Polynesia (5,10), provided estimates of cumulative incidence, but are logistically difficult, require substantial time and resources, and present diagnostic challenges due to varying duration of infection markers (RNA and different types of antibodies) and cross-reactivity (11). Therefore, it is important to find alternative methods to estimate incidence of infection, including statistical techniques that can be easily applied to surveillance data.

Although many infections are undetected during outbreaks, data exist for the set of infections captured through surveillance systems. Bayesian statistical methods explicitly consider both variability in observations (data) and uncertainty in model parameters (e.g., probability of observation), and are well-suited to address challenges like estimating quantities that are not directly observed. During the emergence of ZIKV in Martinique, *Andronico et al*. (12) developed a Bayesian model to explicitly incorporate a classic epidemiological compartmental model with surveillance data from Martinique using prior information on ZIKV transmission, reporting rates, and GBS risk from French Polynesia. We employed a similar approach in Puerto Rico incorporating multiple surveillance indicators and prior information on the probability of observing infections.

We considered surveillance data on suspected arbovirus cases, suspected Zika-associated GBS cases, and infections identified through a subset of blood banks as indicators of infection. Suspected arbovirus cases identified through passive surveillance reflect symptomatic care-seeking individuals with symptoms indicative of ZIKV, dengue virus, or chikungunya virus infection. Suspected Zika-associated GBS cases represented a more severe and easily recognized manifestation, though GBS can also result from other causes. Blood donor data provided information on asymptomatic and pre-symptomatic infections identified through blood screening. To capture underlying infection dynamics, we used a generic Time-series Susceptible-Infected-Recovered epidemic model within a Bayesian framework to relate infections to data by utilizing evidence-based assumptions on detection probabilities for each indicator. In this framework, we estimated the weekly incidence on ZIKV infection and the cumulative number of infections in Puerto Rico in 2016.

## Methods

### Data

We collected data on suspected arboviral disease cases, suspected GBS cases, and infections detected among blood donors reported in Puerto Rico during January 1, 2016–December 31, 2016. A suspected arboviral disease cases was defined as any patient with clinically-suspected illness resulting from an arbovirus infection and reported through the Passive Arboviral Diseases Surveillance System (PADSS) in Puerto Rico (3,13). Suspected GBS cases were patients experiencing onset of neurological symptomatic characteristic of GBS (e.g., bilateral flaccid limb weakness (14)) reported through the GBS Passive Surveillance System–a surveillance system capturing GBS cases along with patients experiencing other neurological symptoms (e.g., encephalitis (14)) and operated by the Puerto Rico Department of Public Health (PRDH) with support from the Centers for Disease Control and Prevention (CDC) (15).

Not all suspected GBS cases were confirmed to be either ZIKV infections or GBS cases, rather they represented real-time reports of possible GBS cases. We used an indicator for asymptomatic and pre-symptomatic infections using blood bank data beginning in April 2016 when all blood donations were tested for ZIKV RNA (16). Two blood collection agencies provided the numbers of total donations and the number testing positive for ZIKV (16), the population of donors was not representative, being predominantly male and not including anyone under age 16. For the population of Puerto Rico, we assumed the population was approximately 3.4 million people based on 2016 census estimates (17).

### Epidemic model

We developed a generalized Bayesian discrete Time-series Susceptible-Infected-Removed model to fit surveillance data over the 52 weeks of 2016 to estimate weekly ZIKV infection incidence in Puerto Rico. We used an underlying SIR epidemic model, where a proportion of the population was infected each week, *z*_*t*_, and was defined as the product of the proportion infected in the previous week (*z*_*t*−1_), the proportion of population that was susceptible in the previous week (*s*_*t*−1_), and a time-varying transmission rate, *β*_*t*_:

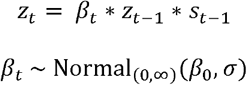

where we use the notation Distribution_(a,b)_(θ) to indicate that we use the stated Distribution with relevant parameter(s) θ but restricted to support (a,b), and so scaled to yield a valid distribution. The time-varying transmission rate (*β*_*t)*_ was assumed to have a constant mean reflecting no substantial control, with random variability between weeks, and was constrained to be greater than or equal to zero using a half-normal distribution. We assumed a prior for the transmission rate (*β*_0_, *β*_0_∼ Normal_(0,∞)_ (2,1)) reflecting an expected weekly reproductive number on the order of 1 to 5 (18). The prior for the standard deviation had a similar scale (*σ* ∼ Normal_(0,∞)_ (0, 1)). Though the average time between successive generations of arboviral infections, i.e., generation times, are typically several weeks (19,20), we implemented this model with a weekly time step, intended to reflect a generic representation of a weekly transmission process in which *β*_*t*_ cannot be directly interpreted as *R*_0_, the basic reproductive number. We did not explicitly aim to model the initial phases of the epidemic, and therefore only estimated an initial proportion of the population infected in the first week of 2016, using a restricted normal prior to indicate a small prevalence of infection that week (*z*_0_∼ Normal_(0,1)_(0, 0.001)). All other individuals were assumed susceptible to infection, as evidence suggests only very limited transmission prior to 2016 (2). Our model assumed a closed population, meaning that susceptible and infection population estimates depended only on population-level risk, and that there were no births, deaths, or migration.

### Reporting models

For each surveillance indicator, we estimated the probability of observing an infection as a function of infection risk (*z*_*t*_) and the observation process for each data type within the epidemic model. The combined model estimated incident infections from the three individual indicators within one epidemic model.

### Suspected arboviral cases

Given that laboratory testing identified very few dengue and chikungunya cases (3), we assumed most suspected arboviral cases were suspected Zika cases. We estimated the expected number of reported suspect arboviral cases as the product of population size (*N*), ZIKV infection prevalence (*z*_*t*_), and the probability of reporting a clinical suspect Zika case per infection (*p*_*S*|*Z*)_ and fit the case data using the negative binomial distribution formulated as a mean and dispersion:

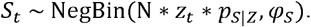

For *p*_*S*|*Z*_, we used a beta-distributed prior to approximate a mean of 0.11 and 95% credible interval [CrI] of 0.01-0.24 based on Mier-y-Teran et al. (21) (*p*_*S*|*Z*_ ∼ Beta(3.3, 27)) (Table 1). The dispersion, *φ*_*S*_, was assigned a prior distribution with high expected overdispersion, *φ*_*S*_ ∼ Normal_(0,∞)_(0, 1000).

**Table 1.** Model parameters and prior distributions.

| Parameter | Description | Mean | 95% CI | Distribution | Reference |
| --- | --- | --- | --- | --- | --- |
| Population (N) | 2016 population for Puerto Rico | 3,400,000 | - | - | (17) |
| Suspect Zika probability ($p_{S\vert Z}$) | Probability of ZIKV infection becoming a suspect case | 0.11 | 0.01, 0.24 | Beta | (18) |
| Viremic detection (D) | Period of detecting Zika virus RNA in blood (days) | 10 | 3, 18 | Weibull | (16) |
| Asymptomatic proportion ($p_A$) | Proportion of ZIKV infections that are asymptomatic | 0.68 | | Beta | (5,6,16) |
| GBS probability ($p_{G\vert Z}$) | Probability of GBS given ZIKV infection | 1.39/100,000 per week | | Beta | (35) |
| Global baseline GBS risk ($p_{G0}$) | Weekly GBS risk due to other causes | 1.89/100,00 per week | | Beta | (21) |

Abbreviations: CI, Confidence interval; GBS, Guillain-Barré Syndrome; ZIKV, Zika virus

### Suspected ZIKV-associated GBS cases

We assumed the number of observed suspected GBS cases (*GBS*_*t*)_ came from a binomial distribution:

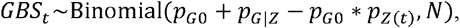

where *p*_*G*0_ is the weekly risk of GBS due to other causes and *p*_*G*|*Z*_ is the probability of suspect GBS given ZIKV infection and *p*_*G*0_ + (*p*_*G*|*Z*_ + *p*_*G*0_ * *p*_*Zt*_) represents the probability of an individual in population N being reported as a suspected GBS case. We assumed the global baseline GBS risk was 0.8-1.9 GBS cases per 100,000 per year (22) for Puerto Rico, and the weekly risk was approximated using a beta prior distribution: *p*_*G*0_ ∼ Beta(23, 8.9 *x* 10^7^). The prior for the probability of suspected GBS given ZIKV infection *p*_G|Z_ was based on an estimated range of 0.5-4.6 GBS cases per 10,000 ZIKV infections (21), which was approximated with a beta distribution (*p*_*G*|*Z*_ ∼ Beta(5.9, 2.3 *x*10^4^)).

### Blood bank indicator

We used an adjustment factor (*f*_*BB*_) to account for the duration of test positivity and the proportion of infected individuals excluded from donating blood because they were symptomatic at the time of donation (16). Assuming weekly testing, the equations and distributions of *Chevalier et al.* were used to approximate a prior distribution for this factor:

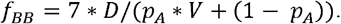

We sampled from distributions of each component, p_A_, the proportion of asymptomatic infections (5,6,11);, the duration of viremia (23); and, the incubation period (23,24), to estimate a Gamma prior for *f*_*BB*_: *f*_*BB*_ ∼ Gamma(6.7, 7.6).

### Analysis of priors

We examined the effect of different prior distribution assumptions on model posteriors for the probability of a suspected case being reported (p_S|Z)_, the probability of acquiring GBS if ZIKV infected (p_G|Z)_, and the relative incidence of ZIKV in the general population compared to positivity in blood donor (*f*_*BB*_) parameters. We assessed the models with three alternative types of prior variance: informative (as described above), informative with doubled standard deviation in the prior and naïve (uniform or flattened). Prior distributions for each parameter under each assumption are available in Supplemental Table 1.

### Model fitting

We fitted epidemic models using a Markov chain Monte Carlo (MCMC) Bayesian framework to estimate incident ZIKV infections and 95% credible intervals [95% CrI] from the three surveillance indicators individually and using all three indicators combined. For each indicator model, we performed 1,001,000 iterations for three chains, and discarded the initial 1,000 iterations as the burn-in period. We evaluated convergence using the Gelman-Rubin diagnostic (25) and thinned the output using every 1,000th sample to obtain 1,000 effectively uncorrelated simulations per chain. For the MCMC simulations, we used the rstan package version 2.19.3 of Stan (version 2.18.0-1, Stan Development Team, http://mc-stan.org), and coda version 0.19-2 (26) package in R version 3.3.2 (https://www.R-project.org/).

## Results

### Estimated infections based on individual surveillance indicators

In 2016, 65,820 suspected arboviral disease cases, 175 suspected GBS cases, and 360 ZIKV-positive blood donors (out of 54,588 tested), were reported in Puerto Rico (Figure 1A-C). We used these data to estimate the weekly and cumulative ZIKV infection incidence in 2016 for each surveillance indicator independently (Figure 1D). Weekly infections estimated from suspected arbovirus cases and suspected GBS cases had similar trends over time, peaking in August 2016 and declining thereafter. Using the blood bank data, estimated ZIKV incidence peaked in June followed by high incidence through August, and declined afterwards. For cumulative incidence estimates based on each of the three indicators, the median estimate was lowest using suspected GBS cases (880,000 infections) and highest using blood bank testing data (960,000 infections), with substantial overlap of credible intervals (Table 2). Estimates based on suspected arbovirus cases had the lowest uncertainty (95% Credible Interval (95% CrI): 630,000 to 1,200,000) and estimates based on suspected GBS cases had the highest (95% CrI: 420,000 to 1,300,000). The estimated proportion of the population infected during the outbreak was similar across the three indicators, with 27% (95% CrI: 19% to 35%) infected using suspected arbovirus case indicator, 26% (95% CrI: 12% to 38%) using the suspected GBS indicator, and 28% (95% CrI: 19% to 37%) using the blood bank indicator.

**Table 2.** Estimated Zika virus infections and 95% credible intervals [CrI] using three surveillance indicators.

| **Surveillance Indicator** | **Cumulative ZIKV infections**<br>**[95% CrI]** | **Proportion of**<br>**ZIKV infections**<br>**[95% CrI]** | **Median incident**<br>**infections per**<br>**10,000 [95% CrI]** | **Peak weekly incident**<br>**infections per 10,000**<br>**people [95% CrI]** | **Week of peak**<br>**incidence**<br>**[95% CrI]** |
| --- | --- | --- | --- | --- | --- |
| **Suspected arboviral**<br>**cases** | 900,000 [630,000, 1,200,000] | 27% [19%, 35%] | 41 [28, 54] | 130 [93, 180] | 33 [32, 34] |
| **Suspected ZIKV-**<br>**associated GBS cases** | 880,000 [420,000, 1,300,000] | 26% [12%, 38%] | 39 [16, 56] | 130 [66, 240] | 32 [28, 38] |
| **Blood bank data** | 960,000 [650,000, 1,300,000] | 28% [19%, 37%] | 35 [22, 48] | 150 [99, 220] | 28 [24, 34] |
| **Combined indicators** | 890,000 [660,000, 1,100,000] | 26% [19%, 33%] | 40 [30, 52] | 130 [97, 170] | 33 [22, 34] |

Median and 95% CrIs are shown from posterior distributions for each surveillance indicator. Peak week refers to the calendar week of the year associated with peak incidence during the epidemic. Abbreviations: CrI, credible interval; ZIKV, Zika virus

**Figure 1.**
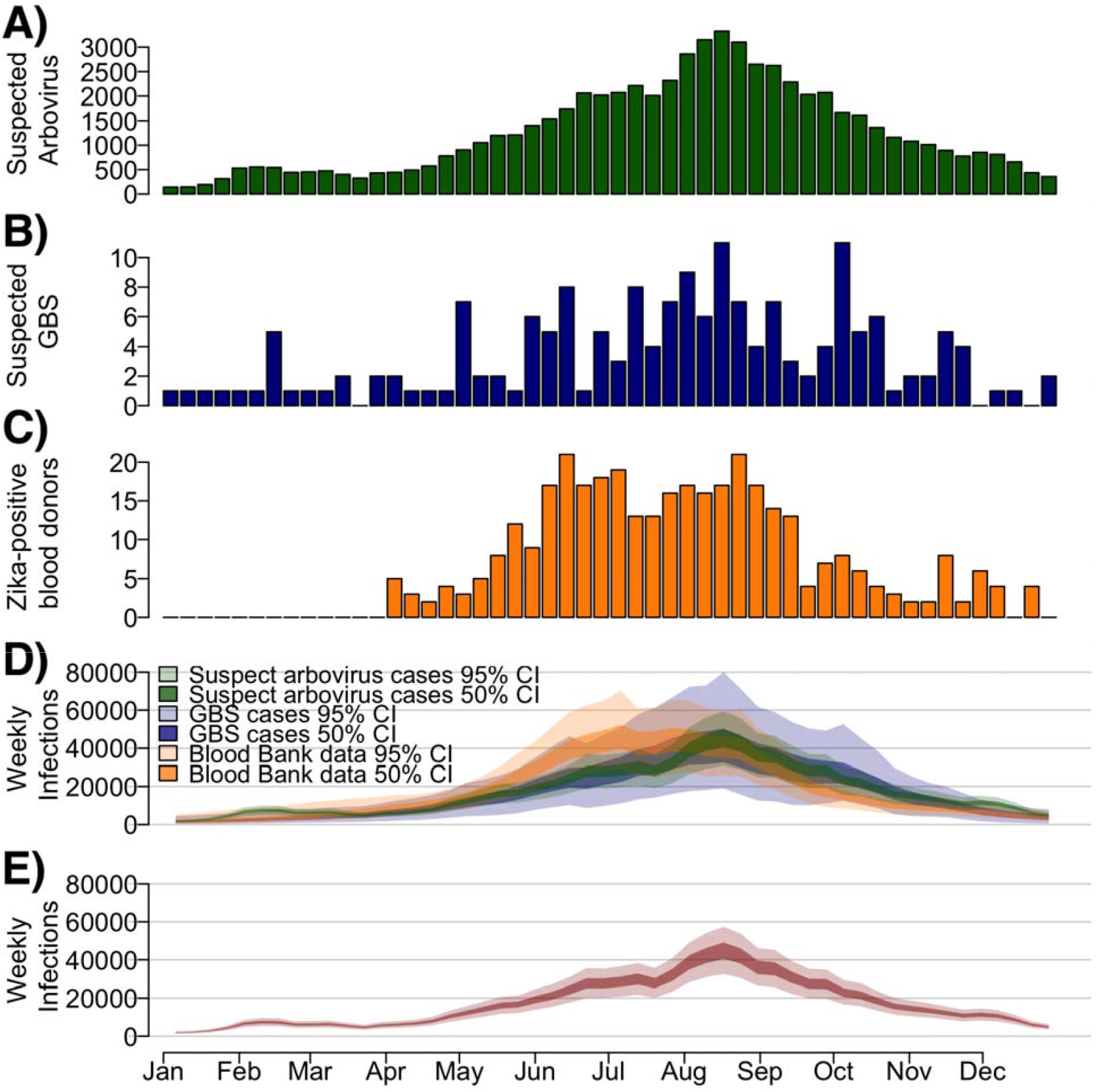
Suspected arbovirus cases, suspected ZIKV-associated Guillain-Barré Syndrome (GBS) cases, ZIKV-positive blood donors, and estimated weekly Zika infections during the 2016 outbreak in Puerto Rico. A) Number of suspected arbovirus cases reported (green). B) Number of suspected ZIKV-associated GBS cases reported. C) Number of ZIKV-positive blood donors identified from blood donor screening. D) Estimated weekly infections using each indicator model separately. Colors refer to each specific indicator used. E) Estimated weekly infections from a model using three combined surveillance indicators. Dark bounds refer to the 50% range (interquartile range) and lighter bounds refer to the 95% credible interval (CrI).

### Estimated infections with combined surveillance indicators

Using a combination of the three surveillance indicators, infections peaked between August and September 2016 (Figure 1E), reflecting the combined peaks in incidence from the three indicators. We estimated that the total incident ZIKV infections was most likely between 810,000 and 970,000 infections (50% CrI, 95% CrI: 660,000 to 1,100,000) (Table 2), corresponding to 24% to 29% (50% CrI, 95% CrI: 19% to 33%) of the total population. These estimates correspond well to an *a priori* triangular probability distribution used to anticipate resource needs during the epidemic (Figure 2) (14,27). The combined estimates had reduced uncertainty compared to that triangle distribution and each independent estimate based on the individual surveillance indicators.

**Figure 2.**
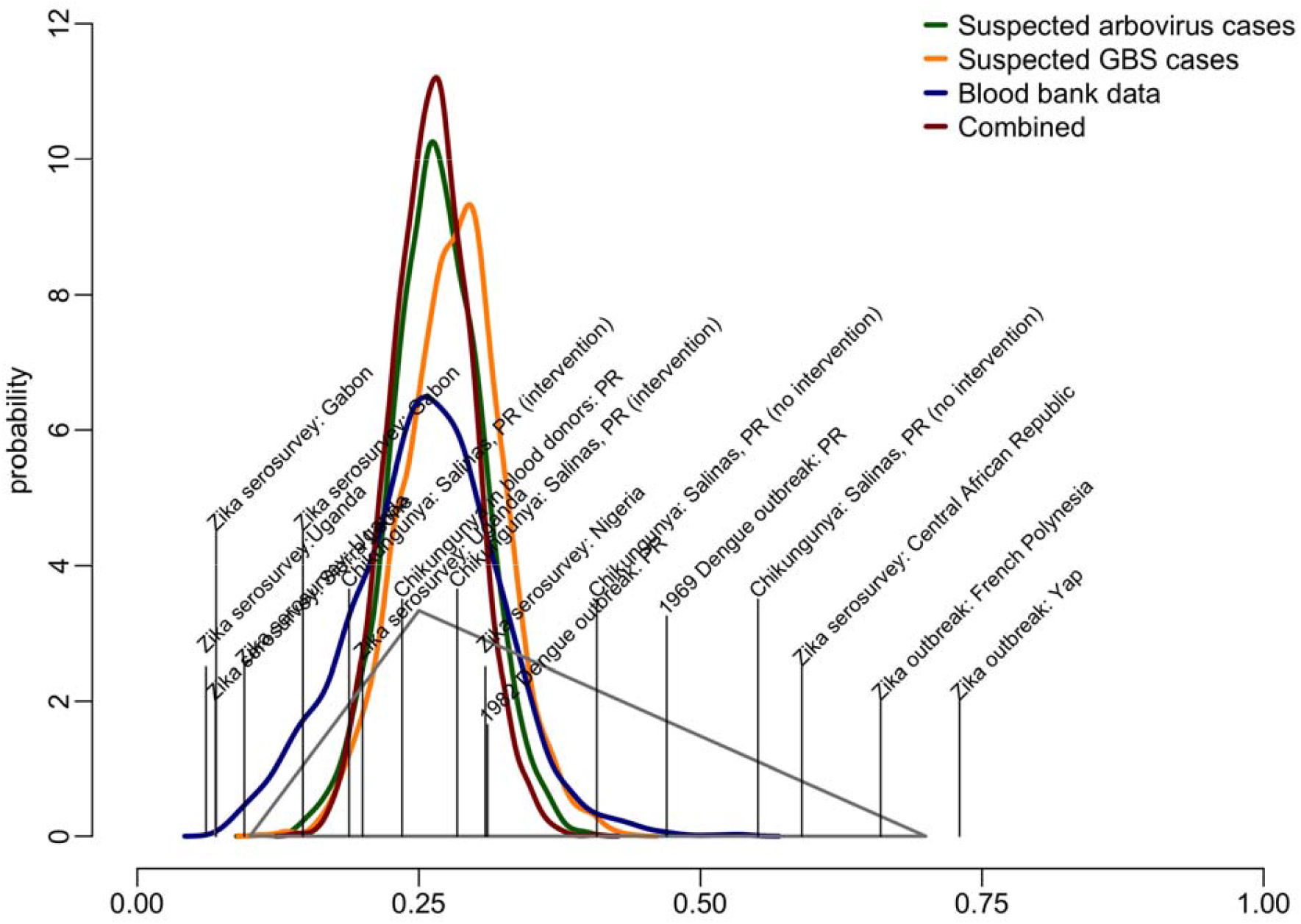
Estimated probability distribution for the proportion of incident ZIKV infections in Puerto Rico in 2016, and probabilities obtained from published literature. The triangle represents the estimated distribution of possible incident infections from *a priori* estimates (15,27) based on previous Zika serosurveys (vertical lines), Zika outbreaks and other arboviral outbreaks in Puerto Rico studies published literature (5,28–31,36–43). Thick lines represent the distribution of the proportion infected estimated from combined surveillance indicators (dark red), and separate surveillance indicators.

### Prior and posterior parameter distributions

For each of the estimates reported above, we used a model with an informed set of prior parameter distributions. However, we also compared these estimates to those with less informed priors (increased variance) and naïve priors (Figure 3A). Increased uncertainty in the priors resulted in similar median estimates for infections throughout 2016, but increased uncertainty especially for the lower bound of the credible intervals (Table 3).

**Table 3.**
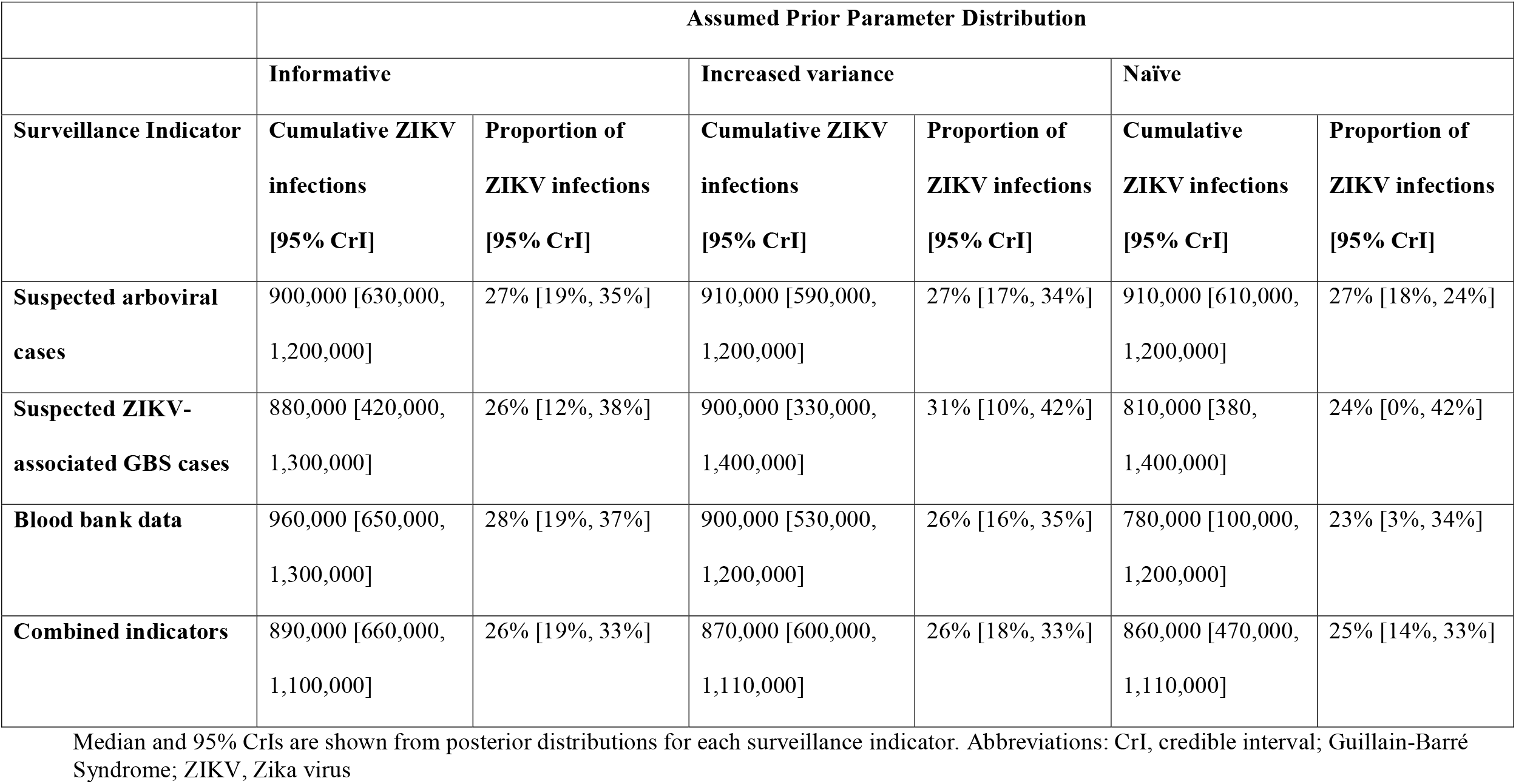
Estimated cumulative Zika virus (ZIKV) infections from models using informative, naïve, and increased variances for prior distributions, Puerto Rico, 2016

**Figure 3.**
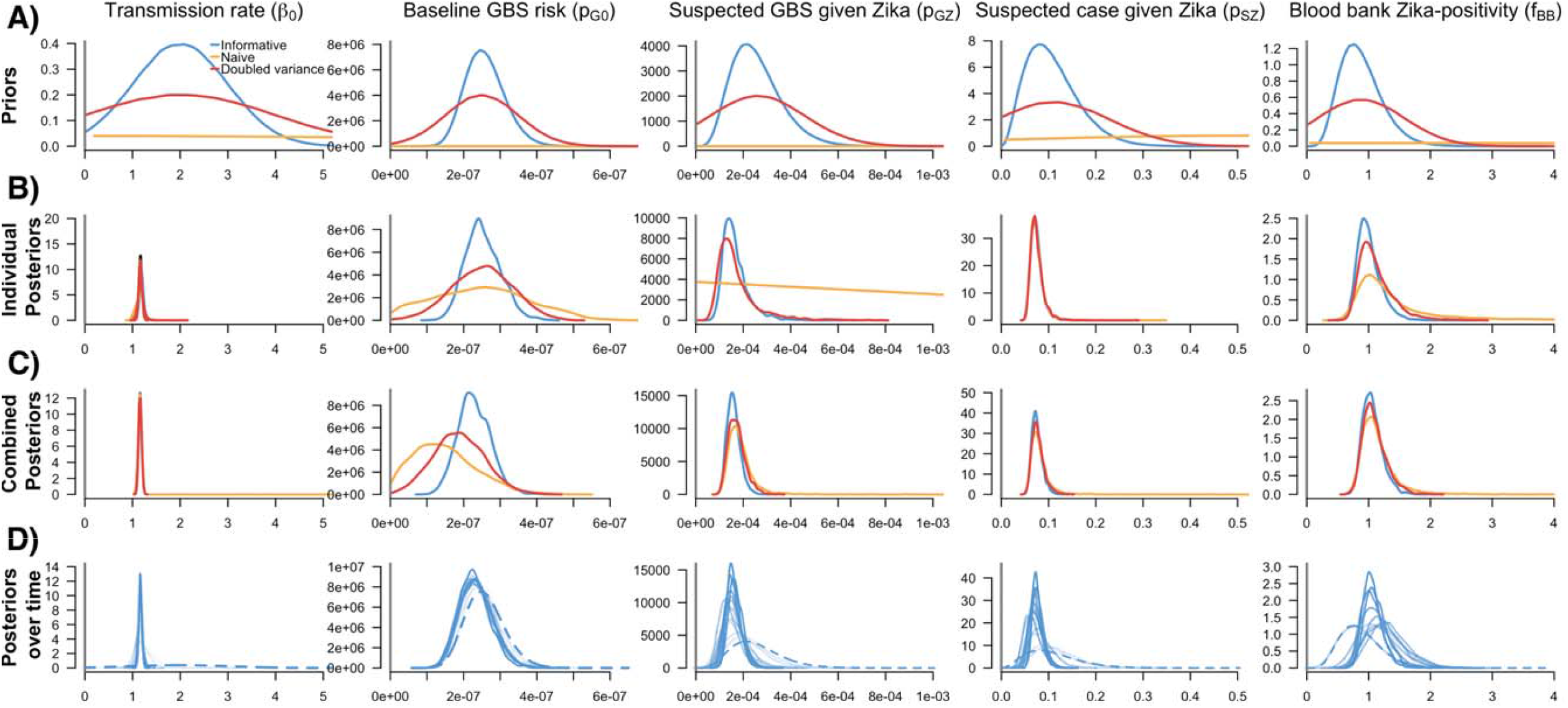
Prior and posterior parameter distributions from individual indicator models, the combined indicator model, and the combined model over time. A) Prior distributions of five model parameters. Color lines refer to assessed variance assumptions of prior distributions in sensitivity analyses. Final individual and combined indicator models used informative priors. B) Posterior distributions of model parameters from individual indicator models. The dashed lines for the Beta parameter refer to the posterior parameter distributions from three models. C) Posterior distributions of model parameters from the combined model. D) Posterior distributions over time (i.e. four-week increments from the end of January 2016 to the end of December 2016) from the combined model using informative priors. Dashed lines refer to the informative priors for each model parameter. Darker transparency of the lines refers to each the posterior distribution from each 4-week increment over time.

For the three prior distributions, the transmission parameter (*β*_0_) converged on a similar value regardless of the surveillance data used. As expected, the less informative priors led to higher uncertainty in the posterior distributions for the outcome parameters, particularly for the individual models in which additional data are not available to inform the estimates (Figure 3B). The most notable effect was seen for the suspected GBS surveillance model. In this case, the posterior baseline GBS risk (*p*_*G*0_) was slightly higher and the ZIKV-specific infection risk (*p*_*G*|*Z*)_ was highly uncertain, indicating a lack of sufficient information to distinguish between the two components of GBS risk. On the other hand, the combined model was able to resolve all parameters regardless of the assumed prior variance. However, parameters using naïve priors still had more uncertainty in their posterior distributions (Figure 3C).

### Posterior estimates over time

Over the progression of the outbreak, the parameter posteriors evolved over time as more data became available for each surveillance indicator (Figure 3D). For the first 4 weeks of 2016, the posterior estimates for individual parameters largely reflected the priors. However, by 8 weeks the posteriors started to shift, narrow, and stabilize.

We observed similar trends when assessing how the incidence estimates of the combined model changed over 4-week increments. When incorporating new data for each surveillance indicator into the combined model over the course of the 2016 epidemic, the incidence estimates had the largest uncertainty in the earliest weeks of the outbreak (Supplemental Figure 1). For each 4-week estimation, the end-of-year estimate based on the full dataset fell within the uncertainty bounds.

## Discussion

Estimates of the true burden of infection for ZIKV, like other pathogens, is challenging because many infections are inapparent; apparent infections are not always recognized, confirmed or reported; and disease surveillance systems for collecting case data are highly varied. Here, we developed an epidemic model, applied within a Bayesian modeling framework, to estimate ZIKV incidence in Puerto Rico using three separate infection indicators available from multiple surveillance systems and assumptions about detection probabilities for each system. Our approach utilizes different data sources to increase the precision of infection estimates over time and may further reduce bias by accounting for inherent surveillance biases based on the probability of detection. Using this framework, we estimated that ZIKV infections occurred in roughly a quarter of the population, resulting in 890,000 total incident ZIKV infections in Puerto Rico in 2016, translating to an average of 36 to 44 new infections per 10,000 people per week. The peak of the epidemic occurred during the week of August 15, 2016 (i.e. week 33), when an estimated 120-140 weekly incident infections occurred per 10,000 people.

We estimated that 19-33% of the population had symptomatic and asymptomatic ZIKV infections in Puerto Rico in 2016, a much higher proportion infected compared to the reported 36,316 confirmed cases (approximately 1% of the population) (3). This estimate was similar to estimates for other arboviral outbreaks in Puerto Rico and to ZIKV estimates generated using other approaches. Community-level dengue seroprevalence studies conducted after emerging outbreaks indicated infection rates of approximately 47% (range: 8-79%, 1969) (28) and 30% (range: 22-45%, 1982) (29). The 2014-2015 chikungunya epidemic resulted in 23.5% seropositivity among 1,031 blood donors in 2015 (30), and for communities participating in a chikungunya vector control study, seroprevalence was 23% (intervention) and 45% (non-intervention) (31). During the ZIKV epidemic, a household-based cluster investigation of 367 participants from 19 clusters found that ZIKV seroprevalence ranged from 0– 57% (4). An estimate based on blood donors alone was 21% (95% confidence interval: 18–24%) (32) while an integrated approach similar to ours estimated an infection rate of 32% (95% CrI: 29-35%)(33).

In contrast to our analysis, studies from other islands found substantially higher seroprevalence estimates, including 73% in Yap in 2007 (5), 49% in French Polynesia in 2013–2014 (10), and 42%–50% in Martinique in 2015 (7,34). The differences in the estimated underlying infection burdens may be in part due to heterogeneity in exposure to infection even within an island population, as seen in early dengue serosurveys in Puerto Rico and municipality-level estimates for ZIKV infection rates (33). Invasion patterns, but nature drive some spatial heterogeneity which can influence longer-term transmission dynamics, since areas with high immunity may limit transmission to areas with low immunity. Other factors that may contribute to differing levels of immunity include human mobility, underlying socioeconomics, and cross-immunity from other arboviruses, and all warrant further examination. The estimated 2016 Puerto Rico infection rate is well below most model estimates for how large a Zika epidemic would be if the population was perfectly mixed (18).

The framework developed here offers advantages beyond the estimation of epidemic size. In practice, we used the model to actively inform situational awareness beginning in August 2016. From the analysis of performance over time, the model could have provided useful information even earlier in the epidemic, despite limited availability of information about the outcome probabilities. Informative priors were available early in the year for baseline GBS risk and for the blood bank reporting factor. While specification of informative prior distributions for the reporting of ZIKV cases and suspected GBS cases would have been more challenging, they would not have been completely naïve. Critically, combining indicators lessened the need for strong prior information for any single indicator. Assuming less precise priors had some effect on outcome precision for all models but had the least effect when indicators were combined, except in the case of completely naïve priors. Similar observations were reported by *Andronico et al.* (12), where examination of different prior assumptions did not strongly affect parameter estimates.

Our approach had some limitations. One of our key indicators, suspected arboviral cases, uses a broad case definition to capture all potential symptomatic cases, and likely includes cases caused by other circulating arboviruses, such as dengue and chikungunya viruses. However, these non-ZIKV cases presumably have had little impact on our indicator, as dengue and chikungunya were rare in Puerto Rico in 2016 (3). We expect if there had been more dengue and chikungunya cases, our framework would improve differentiation from ZIKV cases from other arboviral cases. In general, our prior assumptions were based on data available at the time. If these early data contain hidden bias, resulting estimates could also be biased. Our results suggest that being conservative with respect to precision does not sacrifice substantial precision in the results, in part due to using multiple indicators. Our framework did not allow for substantial transmission variation beyond some week-to-week variability and reduced incidence due to acquired immunity in those already infected. In practice, transmission likely changes seasonally and can be influenced by vector control measures. In this case, with the introduction of a completely novel virus and the lack of large-scale effective vector control measures, we expect there would have been little change in transmissibility throughout the year, but that may not be appropriate in general. Seasonality and other changes in transmissibility can be incorporated into future versions of the framework.

Reported cases generally underestimate the true number of incident infections occurring in an epidemic, since they capture only recognized and reported clinical infections. However, multiple imperfect indicators provide the opportunity to estimate the underlying incidence of infection, utilizing multiple complementary indicators: (1) a broad non-specific indicator with relatively high counts (suspect arboviral disease cases); (2) an indicator of a severe outcome that is rare, but likely to have high reporting fidelity (GBS); and (3) an age- and geography-biased sample of infection prevalence (blood donors). Each indicator offers a unique but biased insight into the progression of the epidemic, capturing different case subgroups including different age-groups, and, when combined, can provide critical situational awareness about the progression of the epidemic.

The approach described here can estimate how many people have been infected in near real-time or identify changes in the trajectory of incidence across various indicators. It is also useful for post-hoc analysis to understand what the impact may have been on the population-level and whether more transmission may be expected. With 19-33% of the population infected in 2016, ZIKV transmission should be much more limited but still possible in Puerto Rico, particularly in areas that may have experienced lower infection rates. These insights are critical both for preparedness for and response to future epidemics, and this modeling approach is applicable to future Zika epidemics as well as epidemics of other pathogens. For instance, using reported influenza-like-illness (ILI)-associated hospitalizations, outpatient ILI visits and reported laboratory-confirmed specimens each have limitations as individual indicators of the incidence of influenza infection, but when combined they may best approximate the incidence of influenza infection. Approaches like the one we present here provide a tool to incorporate these diverse data and the uncertainties in them to generate timely estimates of incident infection and inform response and control efforts.

## Data Availability

All relevant data are within the manuscript and its Supporting Information files.

## Acknowledgements

The findings in this article are those of the authors and do not necessarily represent the official position of the U.S. Centers for Disease Control and Prevention or the U.S. Public Health Service.

## Notes

### Competing Interest Statement

The authors have declared no competing interest.

### Funding Statement

The author(s) received no specific funding for this work.

### Author Declarations

Exemption was obtained from the CDC Human Subjects Research Office as the data were collected as part of regular surveillance activities.

