## Supplementary Information for "Estimating incidence of infection from diverse data sources: Zika virus in Puerto Rico, 2016"

**Supplemental Information**

**Supplemental Table 1. Prior distributions of model parameters using informative, naïve, and increased variances**

| **Parameter** | **Informative** | **Increased variance** | **Naïve** |
| --- | --- | --- | --- |
| Transmission rate ($\beta_{t}$) 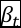 | $\mathrm{Normal}_{\left( 0,\infty\right)}(2, 1)$ | $\mathrm{Normal}_{\left( 0,\infty\right)}(2, 2)$ | $\mathrm{Normal}_{\left( 0,\infty\right)}(0, 10)$ |
| Baseline GBS risk $(p_{G0})$ 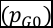 | $\mathrm{Beta}(23, 8.9 x {10}^{7})$ | $\mathrm{Normal}_{\left( 0,\infty\right)}(2.5 x {10}^{-7}, 1 x {10}^{-7})$ | $\mathrm{Normal}_{\left( 0,\infty\right)}\left( 0, 100 \right)$ |
| Probability of Suspected GBS given ZIKV infection $(p_{G\vert Z})$ 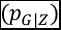 | $\mathrm{Beta}(5.9, 2.3 x {10}^{4}$) 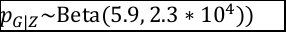 | $\mathrm{Normal}_{\left( 0,\infty\right)}(2.6 x {10}^{-4}, 2 x {10}^{-4})$ | $\mathrm{Normal}_{\left( 0,\infty\right)}(0, 10)$ |
| Probability of a suspected case being reported ($p_{S\vert Z})$ | $\mathrm{Beta}\left( 3.3, 27 \right)$ | $\mathrm{Normal}_{\left( 0,\infty\right)}($0.11, 0.12) | $\mathrm{Normal}_{\left( 0,\infty\right)}(0.5, 0.5)$ |
| Relative incidence of ZIKV in the general population compared to positivity in blood donor ($f_{BB})$ | $\mathrm{Gamma}(6.7, 7.6)$ | $\mathrm{Normal}_{\left( 0,\infty\right)}(0.88, 0.7)$ | $\mathrm{Normal}_{\left( 0,\infty\right)}(0, 10)$ |

Abbreviations: GBS, Guillain-Barré Syndrome; ZIKV, Zika virus

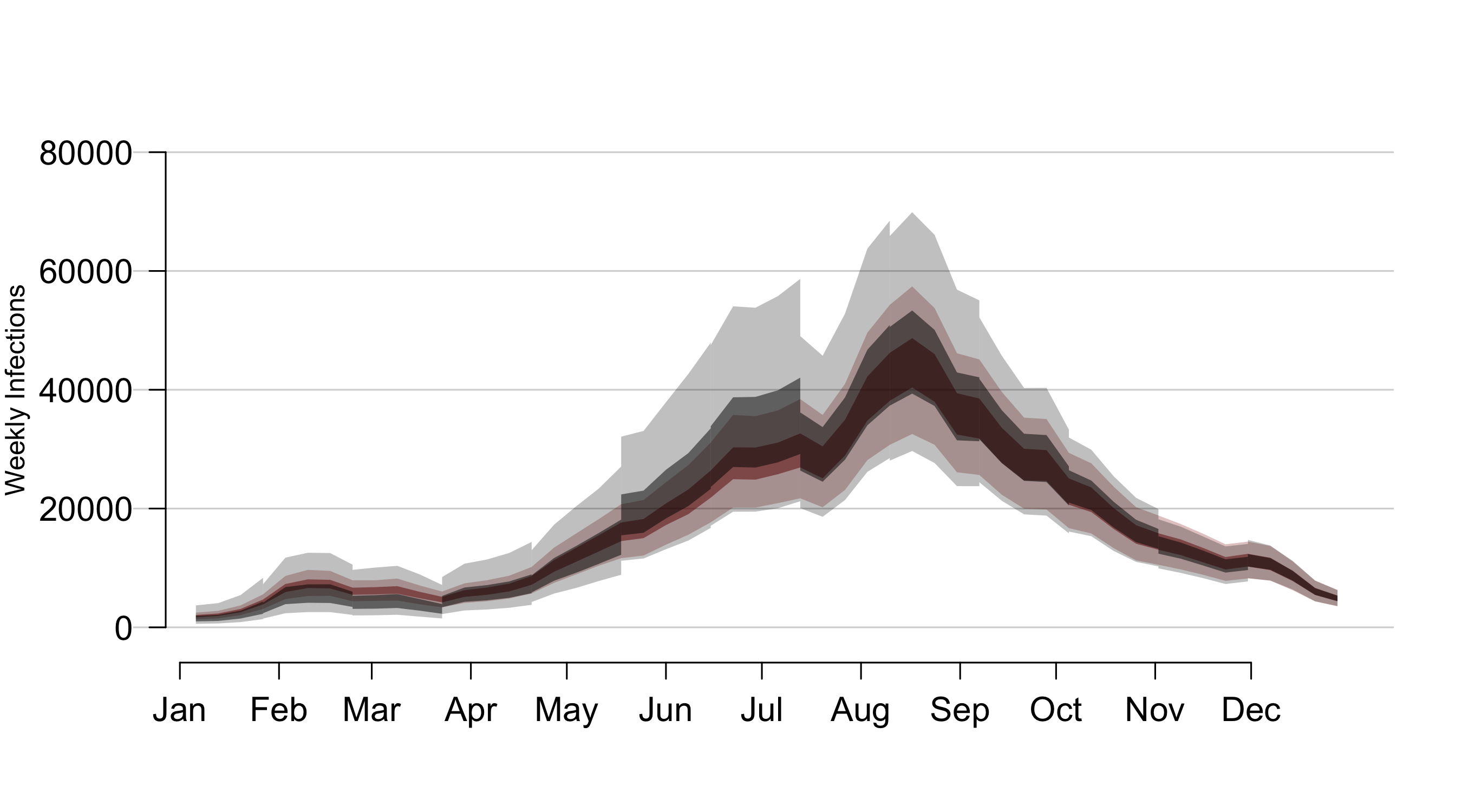
**Supplemental Figure 1. Estimated weekly Zika infections over four-week increments (black) and using all data (red) from the end of January 2016 to the end of December 2016) using the combined indicator model.** Combined models use informed priors. Dark bounds refer to the 50% range (interquartile range) and lighter bounds refer to the 95% credible interval (CrI).
